# Optimization of ^1^H MR spectroscopy methods for large volume acquisition of low concentration downfield resonances at 3T and 7T

**DOI:** 10.1101/2024.04.09.24305552

**Authors:** Neil E. Wilson, Mark A. Elliott, Ravi Prakash Reddy Nanga, Sophia Swago, Walter R. Witschey, Ravinder Reddy

## Abstract

**Purpose:** This goal of this study was to optimize spectrally selective ^1^H MRS methods for large volume acquisition of low concentration metabolites with downfield resonances at 7T and 3T, with particular attention paid to detection of nicotinamide adenine dinucleotide (NAD^+^) and tryptophan.

**Methods:** Spectrally selective excitation was used to avoid magnetization transfer effects with water, and various sinc pulses were compared to a pure-phase E-BURP pulse. Localization using a single slice selective pulse was compared to voxel-based localization that used three orthogonal refocusing pulses, and low bandwidth refocusing pulses were used to take advantage of the chemical shift displacement of water. A technique for water sideband removal was added, and a method of coil channel combination for large volumes was introduced.

**Results:** Proposed methods were compared qualitatively to previously-reported techniques at 7T. Sinc pulses resulted in reduced water signal excitation and improved spectral quality, with a symmetric, low bandwidth-time product pulse performing best. Single slice localization allowed shorter TEs with large volumes, enhancing signal, while low bandwidth slice selective localization greatly reduced the observed water signal. Gradient cycling helped remove water sidebands, and frequency aligning and pruning individual channels narrowed spectral linewidths. High quality brain spectra of NAD^+^ and tryptophan are shown in four subjects at 3T.

**Conclusion:** Improved spectral quality with higher downfield signal, shorter TE, lower nuisance signal, reduced artifacts, and narrower peaks was realized at 7T. These methodological improvements allowed for previously unachievable detection of NAD^+^ and tryptophan in human brain at 3T in under five minutes.

## INTRODUCTION

Metabolites detectable by their downfield protons (>4.7 ppm) such as nicotinamide adenine dinucleotide (NAD^+^), tryptophan (Trp), and nicotinamide riboside (NR) hold significant potential for clinical applications. NAD^+^, a crucial coenzyme in redox reactions, plays a key role in energy metabolism, cell survival, and pathways that modify aging (1,2). Tryptophan, an essential amino acid, is a precursor to serotonin and kynurenine pathway metabolites, implicating its relevance in neuropsychiatric disorders (3–5). Nicotinamide riboside, a NAD^+^ precursor, has potential in treating age-related degenerative diseases due to its role in boosting NAD^+^ levels, thus improving mitochondrial function and cellular resilience (6). These compounds, discernible through advanced ^1^H MRS techniques, offer a promising avenue for understanding and potentially treating a range of medical conditions, from psychiatric and neurodegenerative disorders to aging-related diseases.

There are three main reasons why detection of these metabolites with downfield resonances is challenging. First, their concentrations are lower than the common upfield metabolites that have concentrations in the millimolar range. NAD^+^ has been estimated to be between 200-400 µM in brain with ^31^P (7,8) and downfield ^1^H MRS (9,10) and 290-410 µM/kg wet weight from biopsy of skeletal muscle (11,12). Tryptophan concentrations in postmortem human brain at autopsy have been reported as 20-60 µmol/kg wet weight (13). Second, downfield resonances have often been shown to have shorter T2 relaxation time constants than those found in metabolites with upfield resonances (14–16). Third, a number of downfield peaks experience magnetization transfer effects due to cross relaxation or chemical exchange with water, which reduce their NMR visibility in the spectrum when acquired with conventional spectroscopy sequences utilizing water presaturation (17–19). Taken together, all three factors result in a much lower signal than usually measured with upfield ^1^H MRS.

Specialized sequences have been proposed to combat these issues. Spectrally selectivity excitation (9,19) and metabolite cycling (20,21) have been used to avoid magnetization transfer-related signal loss. Localization has been done with 2D/3D ISIS (15,18,22) and a 1D LASER-analogue (9) to minimize T2 signal decay. Most of these studies have been at ultra-high fields, with only a single study showing detectability of NAD^+^ at 3T (21), and each of these studies have used large volume acquisitions to efficiently enhance SNR. When large volumes are used, regional dependence is lost, and signal can only be described as a global measurement (23). In that case, there is no a priori requirement to localize a voxel as opposed to localizing a slice or even performing unlocalized acquisition in order to minimize TE and increase the excited volume.

Our group has previously used a modified PRESS, triple spin echo sequence with spectrally selective excitation to detect NAD^+^ and Trp in brain as well as NAD^+^ and nicotinamide mononucleotide NMN and NR in skeletal muscle at 7T (10,24,25). This paper looks at optimizing this type of spin echo sequence and its 1D, slice selective analog as well as processing methods for applications in downfield spectroscopy of low signal resonances. Using optimal methods, we show improved spectral quality in brain and calf at 7T compared to our previous work and that NAD^+^ and Trp can be detected in brain at 3T in a scan under five minutes for the first time.

## METHODS

### Acquisition

#### Pulse sequence

A comparison of the proposed sequence to our previous version is shown is Figure 1. As described in more detail below, we compared several spectrally-selective excitation pulses, different localization schemes, outer volume suppression (OVS), and a technique for water sideband removal.

**Figure 1:**
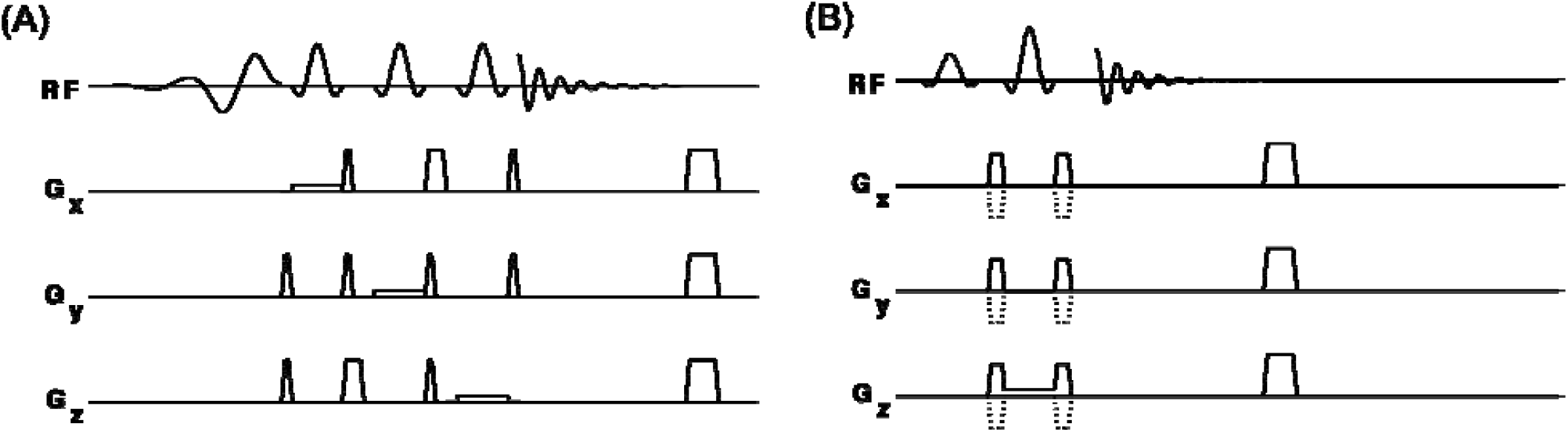
Pulse sequence diagrams of the previous sequence (A) and the proposed sequence (B). The proposed sequence uses a single slice selective spin echo sequence with a sinc pulse for excitation, an SLR pulse for localization, and incorporates gradient cycling of the crushers surrounding the localization pulse. The previous sequence uses a voxel localization formed by the intersection of three orthogonal slices localized by the same low bandwidth SLR refocusing pulse, while excitation used a uniform response, pure phase E-BURP pulse.

#### Localization

Localization was performed in a single shot using spatially-selective spin echoes following excitation. In order to localize a single voxel, three orthogonal slices need to be refocused. Crusher gradient pairs were applied surrounding each refocusing pulse. These crushers had reduced duration and amplitude compared to the vendor-provided sequences in order to reduce the minimum TE without generating eddy current effect. Because of the large voxel sizes and an optimized 16-step phase cycle, no additional, unwanted coherences were generated in the time domain signal (26). This sequence is illustrated in Figure 1A.

Alternatively, as shown in Figure 1B, a single slice can be localized by one spatially selective refocusing pulse, preserving more of the signal when TE is minimized. In this case, a four-step EXORCYCLE (27) was applied over the refocusing pulse, and the crusher gradient pairs are played on all three axes simultaneously. Refocusing pulses in both sequences were 180° symmetric, SLR-optimized pulses (28). At 7T, spectra acquired with relatively low bandwidth (3.1 ppm) refocusing pulses were compared with ones acquired with higher bandwidth (13.3 ppm) which is comparable to that reported with semi-LASER sequences (29).

The relatively narrow bandwidth of the refocusing localization pulses contrasts with conventional spectroscopy sequences where higher bandwidth pulses are typically preferred in order to minimize chemical shift displacement artifact (CSDA) (30). Here, though, we utilized the large chemical shift displacement to shift the localized volume of water away from the brain, further reducing the residual water signal. Figure 2 shows the effective locations of the resulting volumes of interest (VOIs) centered at 9.1 ppm and that of water at 4.7 ppm for a voxel-based localization as well as the difference in volumes over an excitation range of 2 ppm. In the slice-based localization, the water volume is shifted in the slice direction only, while in the voxel-based localization, the water volume is shifted in three directions.

**Figure 2:**
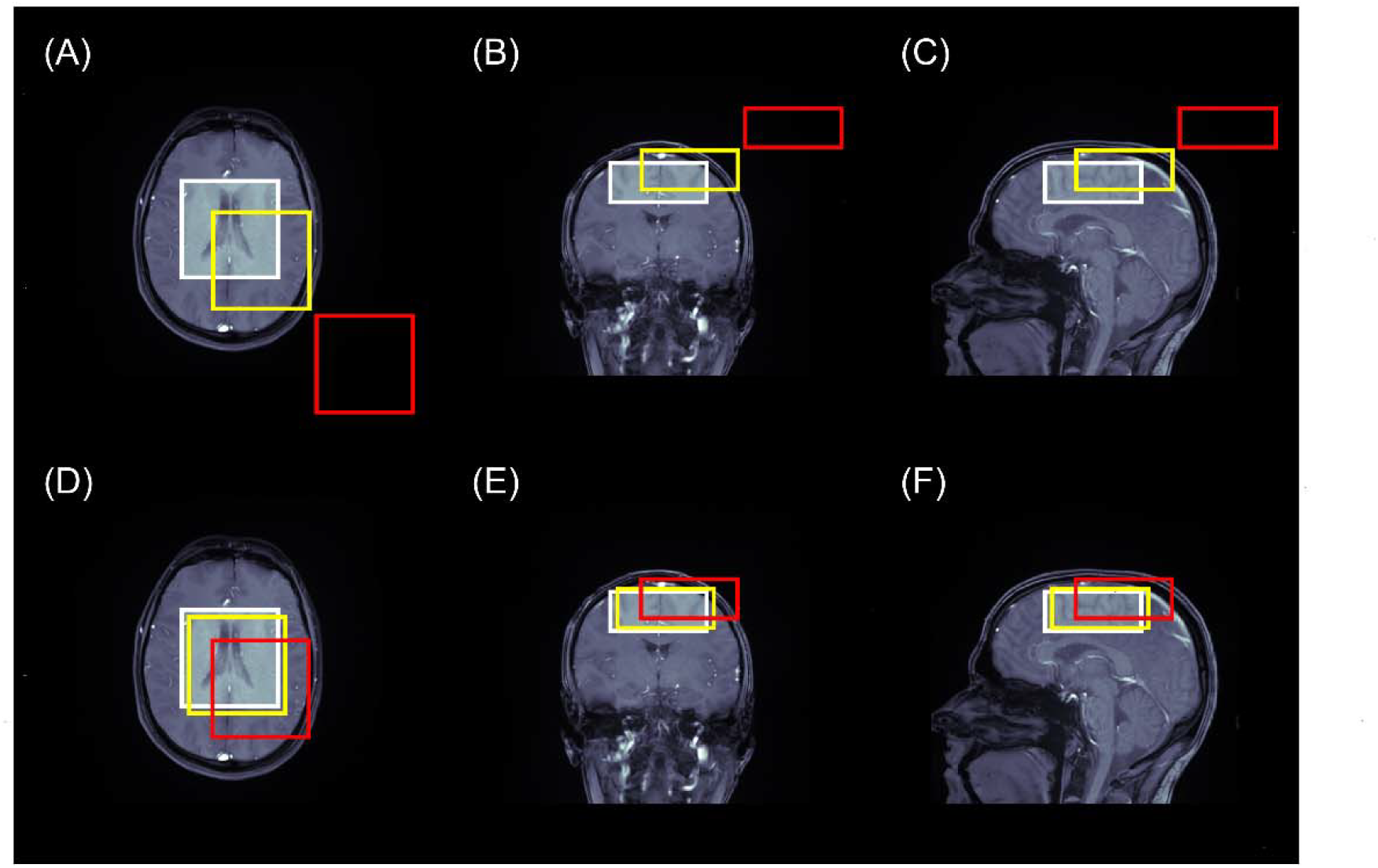
Prescribed VOI of the 9.1 ppm resonance (white box) along with the displaced VOI of water at 4.7 ppm for a typical bandwidth refocusing pulse (yellow box) and for the low bandwidth localization pulses proposed (red box) in the (A) axial, (B) coronal, and (C) sagittal views. (D), (E), and (F) are the same except that the displaced VOI is of the resonance at 8.1 ppm, representing the edge of the desired excitation profile.

#### Excitation

To avoid magnetization transfer effects with water, spectrally selective excitation was employed to leave water signal intact. Here, we compared simple, analytically-derived sinc pulses to a precalculated E-BURP uniform phase excitation pulse (31) that we used in our previous implementations (10,24,25,32). E-BURP does not require any additional refocusing and therefore, does not prolong TE. However, it is defined offline with a single bandwidth-time product and can only be scaled and stretched during runtime. Sinc pulses, on the other hand, can be defined analytically using the following formula for the normalized amplitude

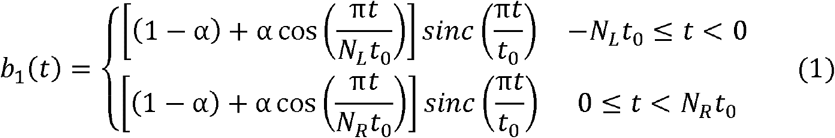

where t_*O*_ is the half width of the central lobe, α is the filter parameter, and *N*_*L*_ and *N*_*R*_ are the number of zero crossings before and after the central maximum, respectively (33). The bandwidth-time product of the pulse is approximated as the total number of zero crossings (*N*_*L*_+ *N*_*R*_). In this study, all excitation pulses had a 2 ppm bandwidth. This resulted in a duration for E-BURP of 7 ms at 7T and 16.3 ms at 3T. Sinc pulses were denoted as Sinc *N*_*L*_ x *N*_*R*_and ranged from having a single main lobe only (Sinc1x1) with duration 3.2 ms at 7T to having three zero crossings on each side symmetrically (Sinc3x3) with duration 24.2 ms at 3T and included Hamming filtering with α = 0.46 in Eq (1). The asymmetric sinc pulses (Sinc4x1 and Sinc4x2) were also tested experimentally and via Bloch simulation.

#### Gradient cycling

Mechanical vibrations of gradient coils can induce time varying magnetic fields resulting in frequency modulation of the signal FID (34–36). Because of the high intensity of the unsuppressed water signal, water sidebands that may appear in the relevant region of the spectrum can be as large as or larger than the metabolite signals of interest. While spectrally selective excitation greatly reduces this artifact compared to broadband spectroscopy without water suppression, there can still be noticeable water sidebands depending on the quality of the signal isolation from water. Since the largest gradient switching in the sequence occurs with the crusher gradients, we applied the gradient cycling technique described by Clayton el. al. (37) in which crusher gradient polarities were reversed every other acquisition. Reversing the gradient polarity causes the sidebands resulting from that gradient to be 180° out of phase so that summation results in significant cancellation of the artifact.

#### Outer volume saturation

Outer volume suppression (OVS) based on slice-selective saturation can be applied around the voxel or slice of interest. We used a modified version of the vendor-provided package in which a 3 kHz bandwidth sinc pulse was played selecting a 2 cm slice followed by a spoiler of moment 19 200 mT/m µs. Options were added to scale the spoiling moment up and to choose the axis or axes the spoiler was played on relative to the saturated slice.

#### Acquisition protocol

This study was acquired under an approved institutional review board protocol at the University of Pennsylvania. All participants gave written, informed consent.

Scans were acquired on a 7T whole body scanner (MAGNETOM Terra, Siemens Healthcare, Erlangen, Germany) and a 3T whole body scanner (MAGNETOM Prisma, Siemens Healthcare, Erlangen, Germany). For brain scans at 7T, a 1 Tx/32 Rx phased array RF head coil (Nova Medical, Wilmington, MA, USA) was used, while at 3T, a similar 32-channel receive only phased array RF coil was used. Calf muscle scans at 7T used a 1 Tx/28 Rx phased array knee RF coil (Quality Electrodynamics, Mayfield Village, OH, USA). T1-weighted anatomical images were acquired for spectroscopic volume positioning. In brain, voxel-localized scans were positioned to cover the superior portion of the head with a thickness of 40 mm, while slice-localized scans were positioned with the same slice center and thickness. The volume was obliqued away from the frontal sinuses for improved B0 field homogeneity. In the calf muscle, volumes were prescribed axially near the most muscular part of the lower leg. Manual shimming was performed utilizing first and second order shims. For voxel-based localization, reported water linewidths were 25-28 Hz, while slice-based localization typically resulted in linewidths 1-3 Hz larger.

Table 1 shows sequence parameters for the different protocols compared here. The minimum TE was used for each protocol unless otherwise noted, while TR was kept fixed at 1 s. Readout consisted of 2048 points acquired with a dwell time of 0.25 ms; 128 averages were acquired for 7T scans in 2 min 12 s (including four preparation scans without readout), while 256 averages were acquired for 3T scans in 4 min 20 s. In each case, a water reference scan was also acquired with excitation centered at 4.7 ppm and averages reduced to the total number of phase and gradient cycling steps with all other parameters remaining the same. Before each readout, the ADC was turned on early with the first four time points discarded to mitigate filter ringing artifacts in the initial part of the FID.

**Table 1:**
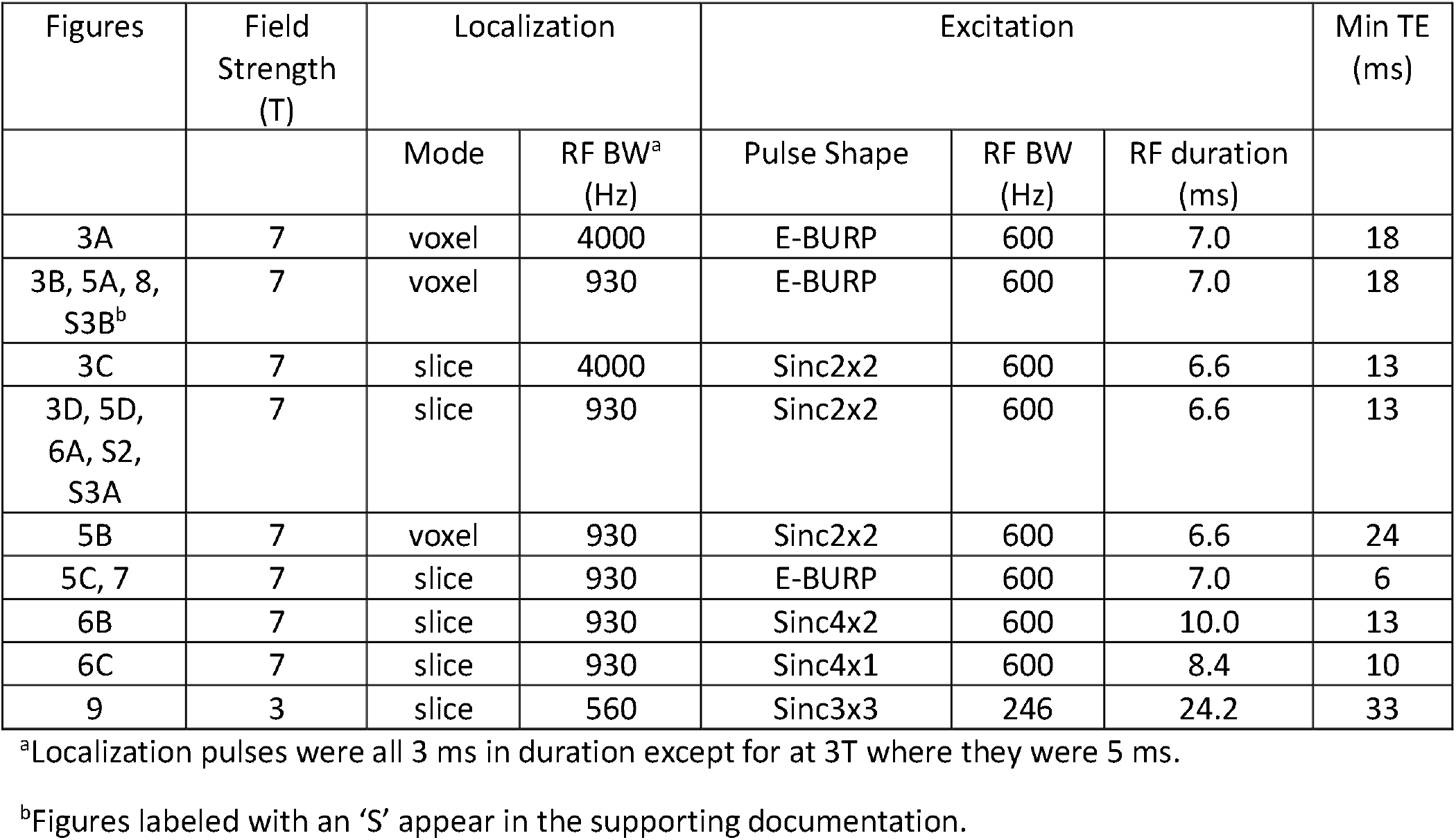
Description of the protocols used in each subfigure.

| Figures | Field Strength (T) | Localization |  | Excitation |  |  | Min TE (ms) |
| --- | --- | --- | --- | --- | --- | --- | --- |
|  |  | Mode | RF BW<sup>a</sup> (Hz) | Pulse Shape | RF BW (Hz) | RF duration (ms) |  |
| 3A | 7 | voxel | 4000 | E-BURP | 600 | 7.0 | 18 |
| 3B, 5A, 8, S3B<sup>b</sup> | 7 | voxel | 930 | E-BURP | 600 | 7.0 | 18 |
| 3C | 7 | slice | 4000 | Sinc2x2 | 600 | 6.6 | 13 |
| 3D, 5D, 6A, S2, S3A | 7 | slice | 930 | Sinc2x2 | 600 | 6.6 | 13 |
| 5B | 7 | voxel | 930 | Sinc2x2 | 600 | 6.6 | 24 |
| 5C, 7 | 7 | slice | 930 | E-BURP | 600 | 7.0 | 6 |
| 6B | 7 | slice | 930 | Sinc4x2 | 600 | 10.0 | 13 |
| 6C | 7 | slice | 930 | Sinc4x1 | 600 | 8.4 | 10 |
| 9 | 3 | slice | 560 | Sinc3x3 | 246 | 24.2 | 33 |

<sup>a</sup>Localization pulses were all 3 ms in duration except for at 3T where they were 5 ms.
<sup>b</sup>Figures labeled with an 'S' appear in the supporting documentation.

## Processing

### Coil combination

As is often recommended, a water signal reference scan was acquired for scaling and coil combination (38). In addition to deriving zero-order phase correction and channel amplitude weighting (39), we used the water reference data to obtain channel-dependent frequency shifts resulting from B_0_ inhomogeneity across large volumes and then applied these shifts to the metabolite spectrum. This was done following a procedure similar to that outlined in Mikkelsen et. al. for robust spectral registration (40,41).

The complex correlation coefficient was computed between the signal from each channel and the remaining channels resulting in a matrix of between-channel *similarity indices*. The median similarity index for each channel was defined as the *similarity* score for that channel. The channel with the highest similarity score was used as a reference signal *fid*_*ref*_. The other channel signals *fid*_*k*_ were then aligned to *fid*_*ref*_ by minimizing the following expression

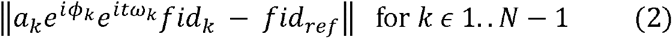

where *α*_*k*_, *ϕ*_*k’*_and *ω*_*k*_ are the fitted amplitude, zero-order phase, and frequency shift for the *kth* channel relative to the reference. Optimization was done using the Levenberg-Marquardt algorithm in Matlab (The Mathworks, Natick, MA, USA). The zero-order phase and frequency shift correction terms were then applied to each water and metabolite FID. An additional pruning step was applied to remove individual channels with low residual similarity scores that fell below a linear regression of the sorted similarity scores after alignment. This process is illustrated for a representative data set in Supporting Information Figure S1.

## RESULTS

### Localization

Slice-based localization was shown to have higher SNR than voxel-based localization because of both the larger signal volume and the reduced echo time. However, this comes at the expense of greater amounts of residual water excitation. Figure 3 shows how using narrow bandwidth localization pulses and sinc excitation helped mitigate the issue. For the wider bandwidth localization pulses, the prescribed voxel at 9.1 ppm and the water voxel had 31% overlap, while the overlap between them was 68% when using slice-based localization. For the narrow bandwidth localization pulses, the overlap is 0% for both voxel-based and slice-based localization. The three NAD^+^ peaks cover a spectral range of 0.4 ppm which has an overlap of 67% with voxel localization and 88% with slice localization.

**Figure 3:**
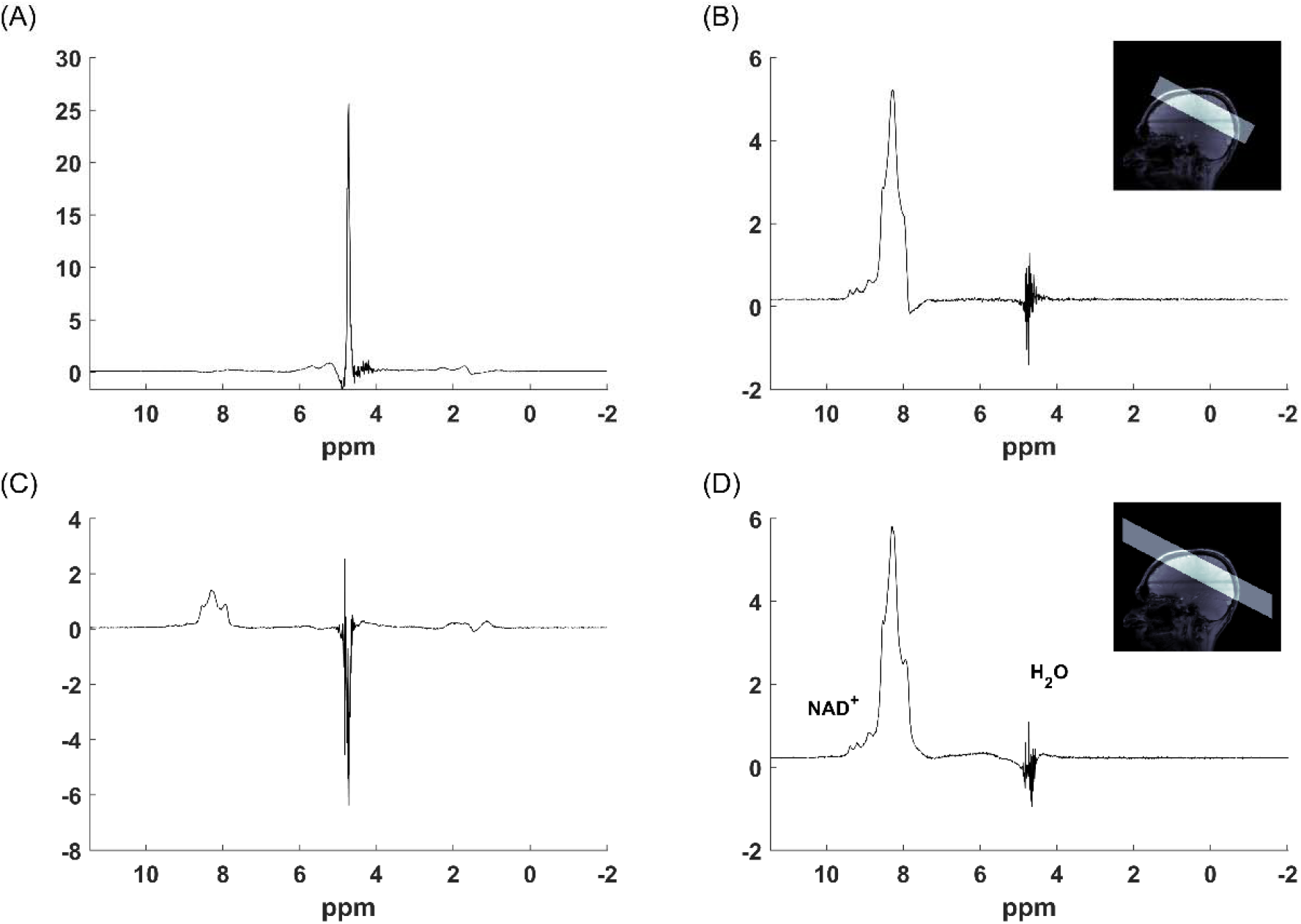
Representative brain spectra from a single session comparing a circumscribed single voxel localization with high bandwidth (A) and low bandwidth (B) localization pulses and single slice localization with the same high (C) and low (D) bandwidth localization pulses. Note that each spectrum is scaled based on its range and that the spectra with high bandwidth localization show significantly higher water signal.

### Excitation

Figure 4 shows the waveform and simulated excitation profiles of the E-BURP pulse as well as various sinc pulses tested. For the E-BURP pulse, readout immediately after excitation was simulated, while for the sinc pulses, an isodelay equal to the time from waveform peak to its end was included before readout to remove the linear phase profile across the spectrum. With the sinc pulses, the width of the transition bands is notably dependent on the number of zero crossings, while the E-BURP pulse shows significantly greater rippling in both the stopband and the passband. This results in reduced water excitation when sinc pulses are used, and this effect is heightened when doing a full slice localization compared to a voxel localization as shown in calf muscle in Figure 5, where Figures 5A and 5B compare voxel-localized spectra acquired with E-BURP and Sinc2x2 excitation pulses, respectively. Figures 5C and 5D compare single slice localized spectra with the same pulses. Note the visibly higher SNR in the slice localized spectrum in Figure 5D compared to that of the voxel-localized spectrum in Figure 5B mainly due to its significantly larger volume of interest.

**Figure 4:**
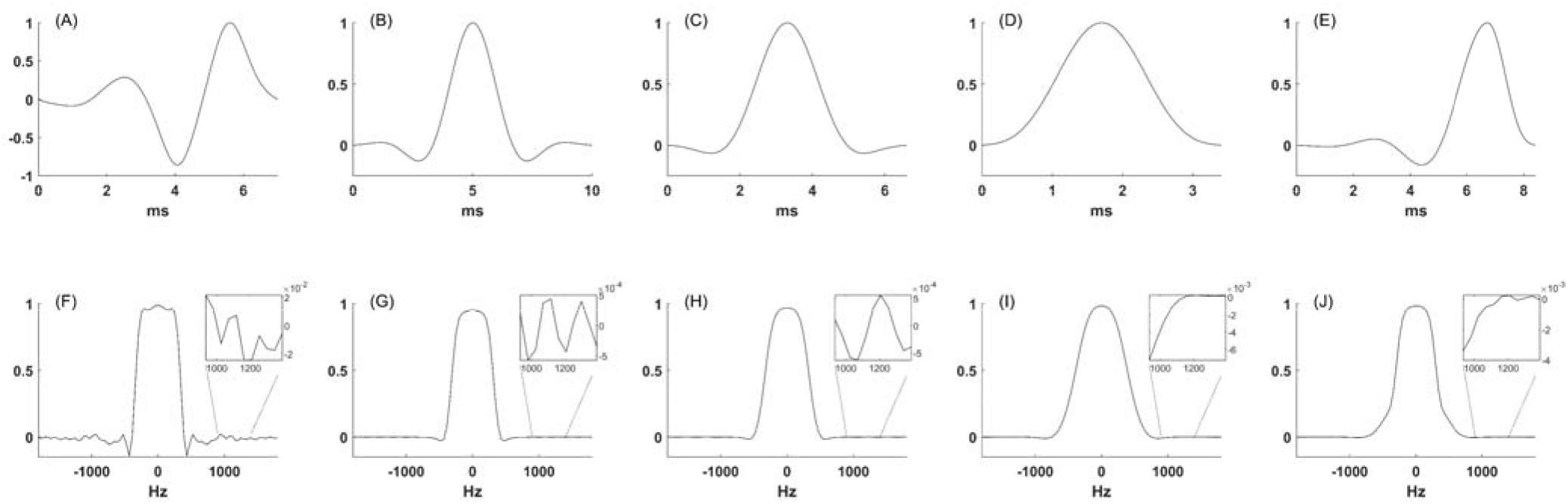
Pulse waveform (top) and simulated spectral excitation profiles (bottom) for E-BURP (A,F), Sinc3x3 (B,G), Sinc2x2 (C,H), Sinc1x1 (D,I), and Sinc4x1 (E,J). Sinc pulses are named based on the number of zero crossing on each side of the central maximum (e.g. Sinc4x2 has four zero crossing before and two zero crossings after the central maximum). Insets show the spectral range around water at 7T for each profile and are scaled individually.

**Figure 5:**
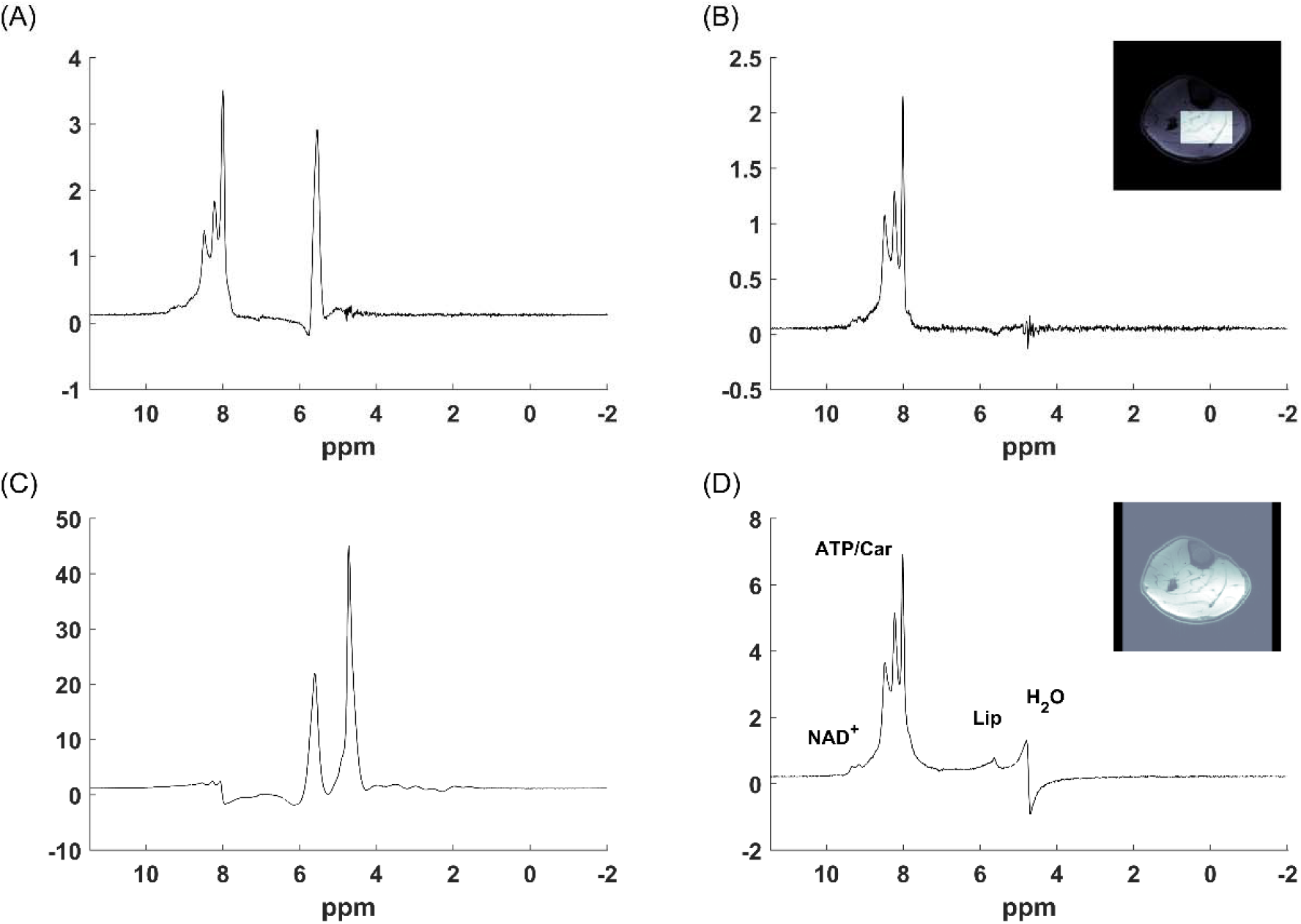
Representative calf spectra with different excitation pulses and localization schemes. An inscribed voxel-based localization with (A) E-BURP and (B) Sinc2x2 excitation. Single slice localization with (C) E-BURP and (D) Sinc2x2 excitation. Spectra were acquired in a single session. Note that the spectra are separately scaled to their respective ranges.

For the applications presented here at 7T, there was not a strong motivation to use sinc pulses with narrower transition bands at the expense of pulse duration because of the large spectral dispersion between NAD^+^ and water/lipids. Figure 6 shows the results of testing asymmetric sinc pulses compared to a symmetric one. The pulses in Figures 6A and 6B have the same isodelay and therefore minimum TE. However, the symmetric pulse used in Figure 6A shows a bit better quality spectrum with more resolvable NAD^+^ and NAA peaks. The highly asymmetric pulse used in Figure 6C shows higher signal but less resolvable peaks and more pronounced phase artifacts at the transition band.

**Figure 6:**
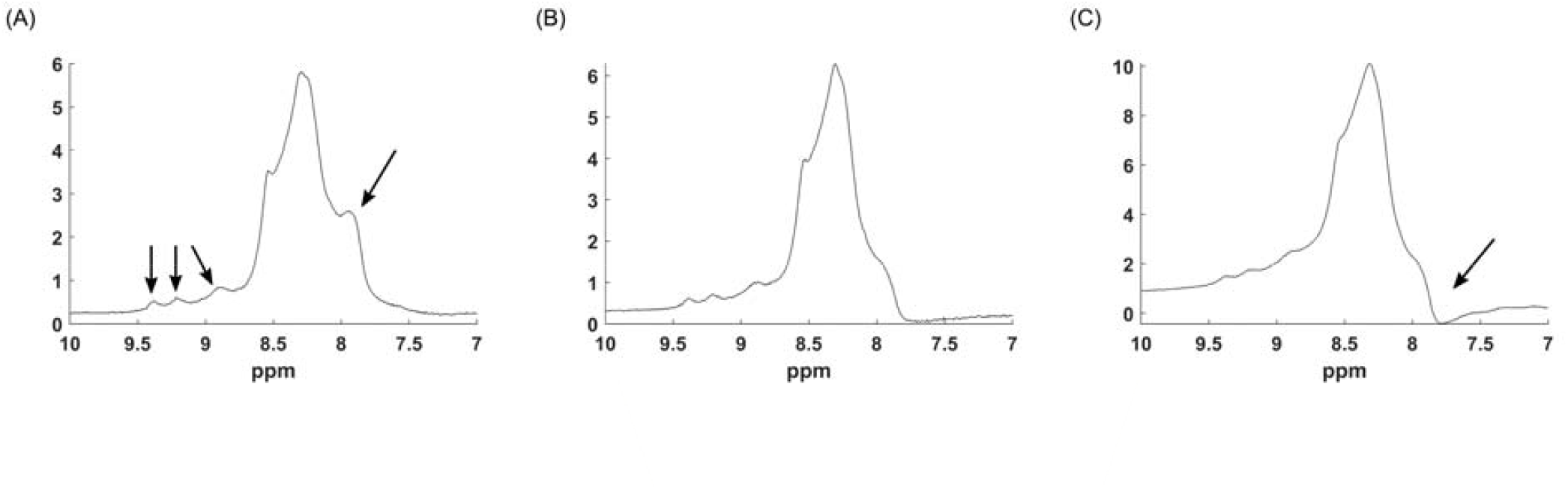
Comparison of symmetric and asymmetric sinc excitation pulse waveforms. (A) Symmetric Sinc2x2, (B) asymmetric Sinc4x2, and (C) strongly asymmetric Sinc4x1. TE was held constant at 13 ms between scans, though the Sinc4x1 pulse has a shorter minimum TE of 10 ms. Arrows highlight the improved peak resolution in (A) and the phase artifact in (C).

### Gradient cycling

As sideband artifacts are proportional to the excited water signal, their appearance can be inconsistent from experiment-to-experiment, depending on several factors including shimming and volume location/orientation, and they can be reduced by focusing primarily on reducing water signal excitation. However, in cases where the excited water signal was sufficiently large, appreciable sideband artifacts are observed, and the use of gradient cycling reduced artifacts in the range where NAD^+^ and Trp resonate as shown in Figure 7. Empirically, we have noted that while sideband artifacts are not always prevalent, gradient cycling has no adverse effect on the spectral quality, and therefore, the use of gradient cycling is recommended. In the sequence, gradient cycling can be nested inside or outside the phase cycling loop (preferred) or as part of the phase cycling loop if the number of acquisitions is limited.

**Figure 7:**
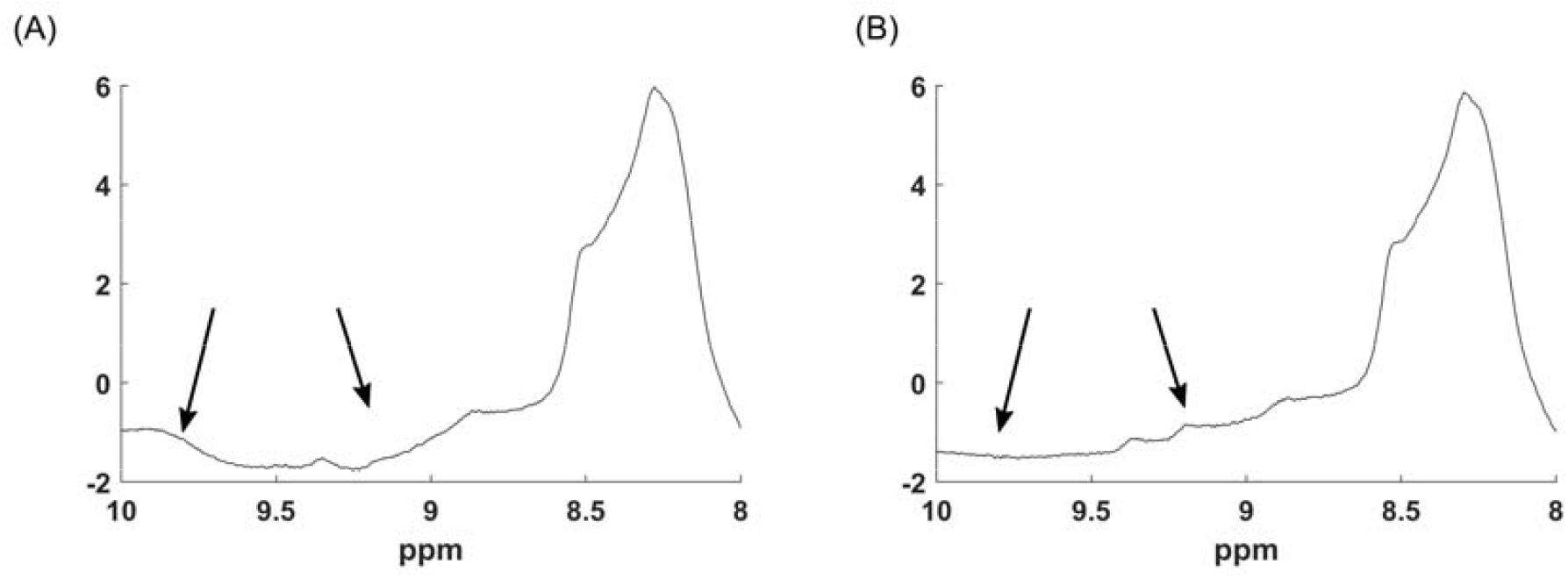
Brain spectra acquired in the same session (A) without and (B) with gradient cycling that large residual water contamination. Other acquisition parameters were identical. Arrows highlight regions where water sidebands obscure the metabolite signal of NAD^+^.

### Outer volume saturation

A consequence of the low bandwidth-time product of the localization RF pulses is that the selected volume does not have sharp edges. In principle, this could be improved using OVS, as can signal reduction from undesired, out-of-volume tissues (e.g. lipids from the skull or subcutaneous fat in skeletal muscle). However, in practice, when following the protocols in this paper, the outer volume artifacts caused by water and lipid are benign in the downfield region of interest, and the addition of OVS actually increased the amount of nuisance signal excitation despite some effort to optimize the module (results not shown). This is most likely due to the unintentional refocusing of unwanted coherences, but further optimization of the OVS module is beyond the scope of this work.

### Coil combination

Individual channel spectra from a representative water reference scan are shown without correction (Figure 8A), after frequency alignment (Figure 8B), and after channel pruning (Figure 8C). Before frequency alignment, zero-order phase was corrected using the phase of the first time point of the fid. However, there were still remaining lineshape differences and center frequency shifts between the individual channels. Most individual channel spectra could be sufficiently corrected using Equation 2, and those that could not were pruned (see Supporting Information Figure S1 for an example). Applying the frequency and phase corrections from the fit of the water signal resulted in higher signal and narrower peaks in both the coil-combined water and metabolite spectra, shown in Figs 8D and 8E, respectively.

**Figure 8:**
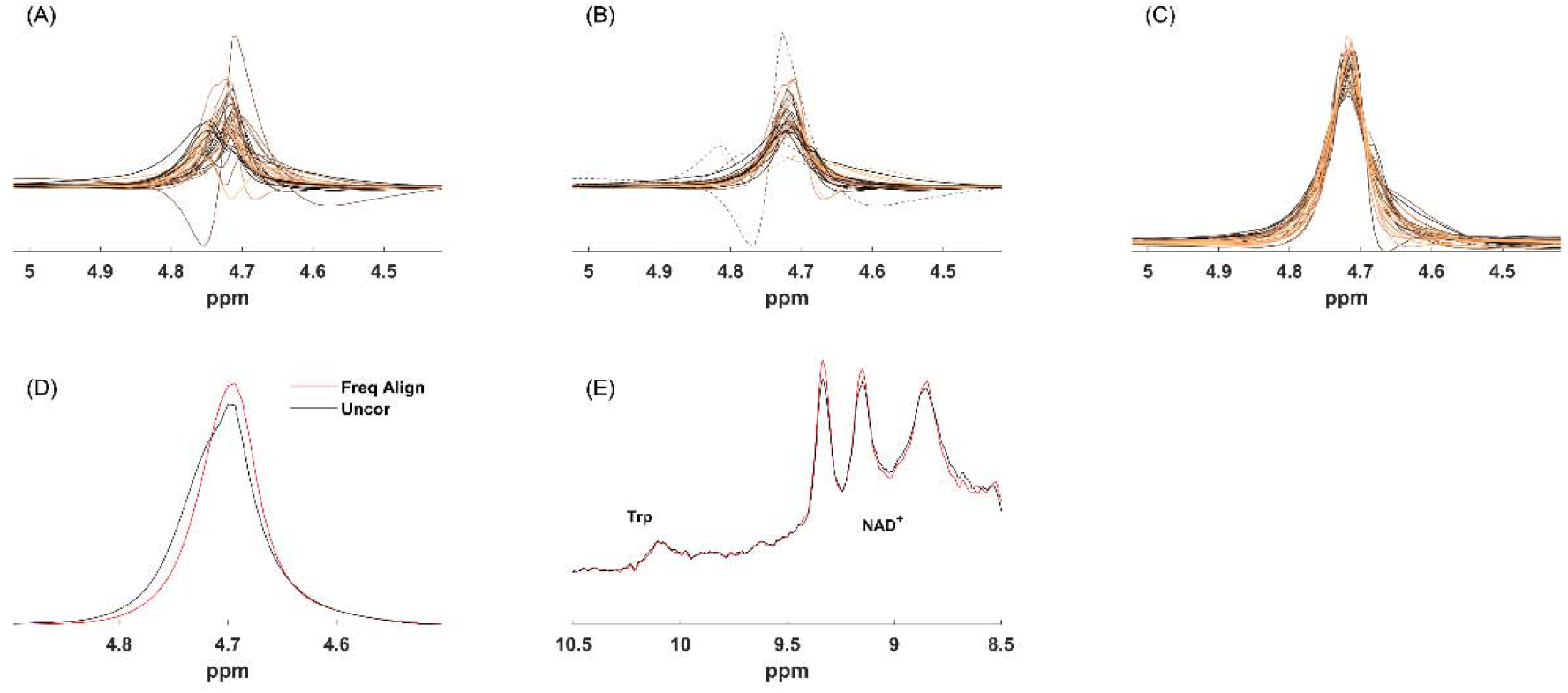
The effect of optimal coil combination. Individual channel water signals (A) without correction, (B) after frequency alignment, and (C) after frequency alignment and low-similarity channel pruning. Coil combined signal of (D) water and (E) metabolites with optimal combination (red) and with standard coil combination (blue).

### 3T spectra

Using an optimized, slice selective version of the sequence shown in Fig 1B with a Sinc3x3 excitation pulse and an extra narrow bandwidth localizing refocusing pulse, we were able to acquire spectra at 3T that show clear and consistent NAD^+^ and Trp peaks from human brain in four different subjects as shown in Figure 9.

**Figure 9:**
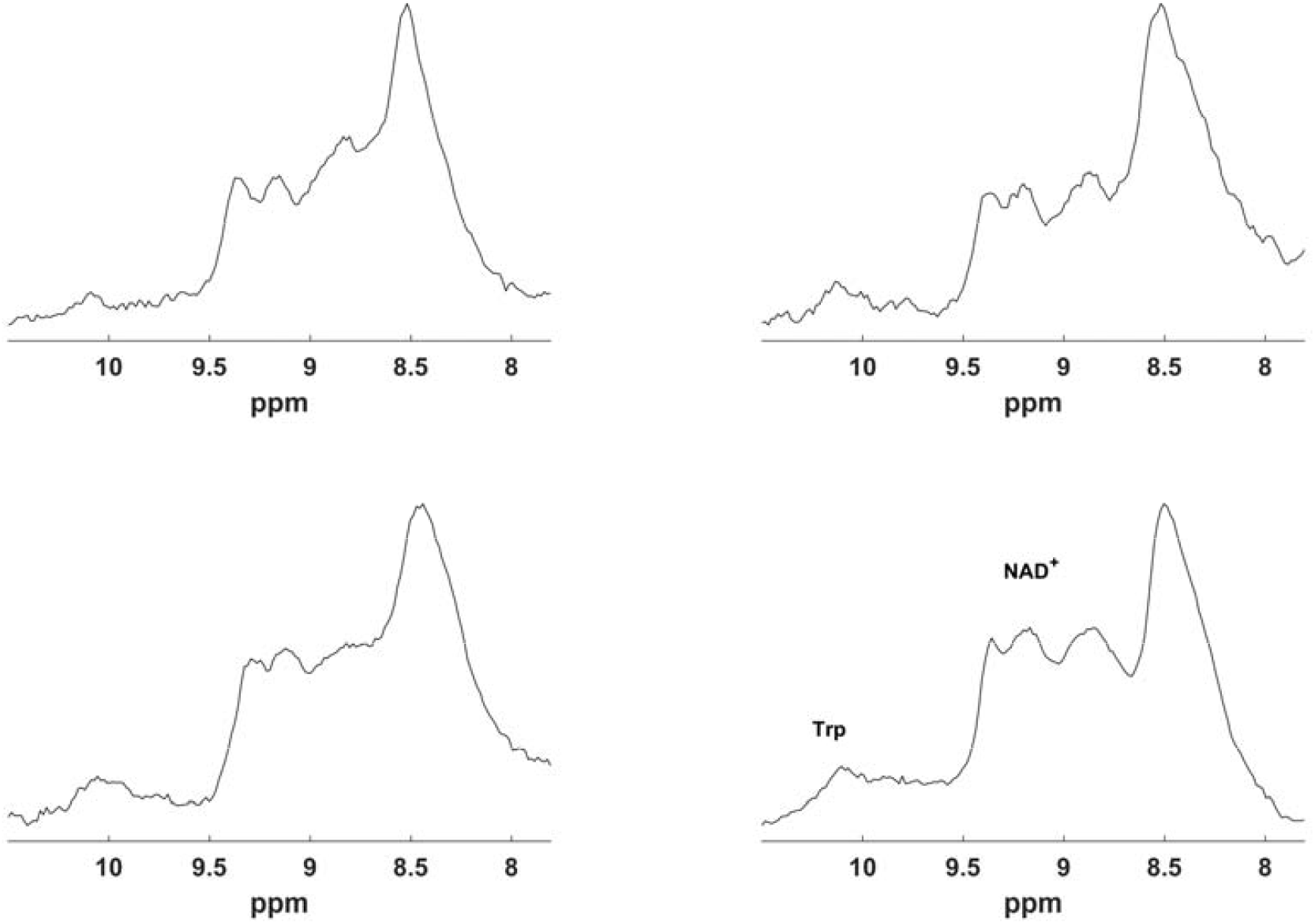
Downfield spectra acquired at 3T showing NAD^+^ and Trp in four subjects using the proposed, slice selective sequence and processing modifications. A Sinc3x3 pulse with 246 Hz bandwidth was used for excitation and centered on 9.7 ppm. Each spectrum was acquired in 4 min 20 s.

## DISCUSSION

In this study, we have taken a component-wise look at the family of spin-echo-based pulse sequences and subsequent data processing for large volume spectroscopy of downfield metabolites in vivo. By looking at these sequences modularly, we better understand how to construct an optimal protocol for a given application. Here, with our attention focused on the detection of NAD^+^, we were able to create optimized protocols at both 7T and 3T for human brain and at 7T for human calf muscle by modifying the waveforms and bandwidths of the excitation and localization pulses as well as the center frequency of excitation.

In upfield ^1^H spectroscopy of human brain, 3D-localized voxel scans are often preferred in order to avoid significant skull lipid artifacts that make quantification of certain metabolites (e.g. lactate) difficult. The same is true in calf muscle where overabundance of subcutaneous fat can obscure intra-myocellular lipid measurement (42). However, in downfield spectroscopy with spectrally selective excitation, minimal lipid signal from the methyl or methylene groups is excited, and signal from the downfield methyne group is relatively small and isolated spectrally. Hence, localization that includes large lipid-containing regions is possible. We have also experimented with unlocalized acquisitions (results not shown) but found that spectral quality is significantly degraded due to difficulty shimming over such a large region. Therefore, we opted to use a single-slice localization where shimming results in only mildly broader linewidths compared to a similarly sized 3D-localized voxel.

In moving from a pure phase selective E-BURP pulse to a traditional sinc pulse for spectrally selective excitation, the greatest drawback is the prolongation of TE to accommodate both the isodelay of the sinc pulse and an additional delay to refocus the linear phase across the spectrum. With spatially selective pulses, the time required to refocus this phase accumulation depends on the available gradient strengths and can be much shorter than the isodelay of the pulse. However, for a spectrally selective pulse, refocusing can only be achieved by waiting the full isodelay time of the pulse, which for sinc pulses is the time from the waveform maximum until its end. This means that the minimum TE increases by twice the isodelay when switching from E-BURP to a sinc pulse. For narrow band excitation, this can be quite large depending on the bandwidth-time product of the pulse. Since the spectral region we are interested in is well separated from water, we have sacrificed a sharp transition band of the sinc pulse in order to keep the isodelay shorter. However, we did find that a sharper transition band was required for 3T compared to 7T due to the reduced spectral dispersion. For a protocol targeting metabolites closer to water, this sacrifice may not be possible, and the increase of the minimum TE might be more severe. In that case, strongly asymmetric pulses should be investigated more thoroughly.

As an alternative to sinc waveforms, spectrally selective SLR pulses could be used and might be superior for certain applications. Here, we chose instead to use sinc pulses for two main reasons. The first is the ease of implementation; sinc pulses can be defined analytically with a simple formula (Eq 1) that readily accommodates changes in bandwidth-time product, asymmetry, and filtering. This can be especially useful when designing new protocols on the scanner. The second reason is the that the excitation profile flatness of the sinc pulse is more robust to B1+ variation than those of E-BURP or SLR pulses which experience larger passband and stopband ripples. B1+ inhomogeneity is particularly problematic at ultrahigh field, especially with large volumes, but it may not be as significant an issue at lower field strengths.

One consequence of the increased CSDA caused by low bandwidth localization pulses is that the direction of the displacement becomes significant. In the head, gradient polarity is chosen such that the water and lipid volumes are shifted superiorly away from the head in the slice direction. In the calf muscle scans, the slice selective gradient polarity was reversed to shift the water and lipid volumes inferiorly towards the foot where leg volume is reduced. As voxel-based localization uses three orthogonal refocusing pulses, the CSDA is experienced in all three directions simultaneously, creating a larger overall separation and lessening the dependence on direction. Supporting Information Figure S2 shows the differences in the water signals in a slice-based acquisition in human brain for alternate polarities of the slice selection gradient.

Our gradient cycling technique is only applied to the crusher gradients surrounding the slice-selective refocusing pulse(s). While it is true that this technique only removes water sidebands originating from vibrations caused by the crusher gradients and that more robust sideband removal techniques exist (36),water sidebands originating from slice selective gradients are expected to be negligible in the present context for two reasons. First, spectrally selective excitation greatly decreases the residual water signal compared to broad band excitation techniques without water suppression. Second, the low bandwidth-time product RF pulses and thick slices mean that the slice selective gradients are lower in amplitude than those typically used in imaging or even conventional spectroscopy. As shown in Supporting Information Figure S2, for low bandwidth refocusing, reversing the polarity of the slice selective gradient in order to cancel water sidebands is not feasible as entirely different volumes with different water content are localized.

In addition to causing spectral sidebands, pulsed gradients can also generate eddy currents in which a time dependent magnetic field causes spurious peaks and lineshape distortions. Again, crusher and spoiler gradients contribute most to this phenomenon. The sequences presented here have reduced crusher amplitudes surrounding refocusing pulses compared to standard, vendor-provided single voxel sequences. In addition, because of the spectrally selective excitation, there are no spoiler gradients as part of a water presaturation module, which is a significant source of eddy currents in conventional ^1^H spectroscopy. Therefore, we do not expect eddy currents to be a significant source of artifacts in these sequences, and this is supported by each peak of interest being well fit to a single Lorentzian in the HSVD algorithm (43).

The most common method of eddy current correction (ECC) involves collecting a pure, on resonance water signal and subtracting off its time domain phase from the metabolite signal (44). A consequence is that both the zero order phase and frequency shifts of water are removed, similar to our proposed method of coil channel combination based on spectral registration. However, there are a couple of important differences between the two approaches. First, since ECC is ostensibly used to mitigate time dependent phase changes, each time point in the FID is corrected individually with the corresponding point in the water reference FID. In contrast, spectral registration-based correction uses hundreds of points to solve for only three parameters making it more robust and accurate when time varying eddy current signals are not present. Second, the proposed method includes a procedure for pruning channels that cannot be well corrected and do not contribute coherently to the signal, while ECC does not. Pruning has been shown to improve spectral quality in large volume spectroscopy (23). Nevertheless, ECC is a viable method to perform channel-wise spectral alignment for large volume spectroscopy, and a comparison with the proposed method is shown in Supporting Information Figure S3.

Because of the reduced spectral dispersion at 3T, residual water signal is a more significant nuisance than at 7T. Therefore, methods specifically focused on reducing residual water signal were most effective. This included extra narrow bandwidth localization pulses set to mimic the CSDA present at 7T as well as sinc spectrally selective excitation. The bandwidth of the excitation pulse was reduced, its center frequency was shifted further downfield, and the number of zero crossings was increased to sharpen the transition band. Together, these changes reduced residual water signal sufficiently, where the downfield metabolites were readily identifiable. However, a consequence of these changes is that the minimum TE increased from 13 ms at 7T to 33 ms at 3T with a single slice localization. Voxel-based localization with the same pulses has a minimum TE of 48 ms which is prohibitively long for short T2 downfield metabolites. Despite the reduced signal, spectral SNR was still high enough using the protocol shown here to observe NAD^+^ and Trp peaks in a less than 5 minute scan, marking the first observation of Trp in the ^1^H spectrum at 3T and a significant acceleration compared to the 32 minute protocol in Dziadosz and Hoefemann et. al. for detection of NAD^+^ (21).

A major limitation of this study is that the signal from such large volumes can only be considered a global representation and is not sensitive to regional differences. Recent work using MRSI at 3T has shown promise in obtaining downfield spectra from smaller volumes, though it has currently been restricted to larger signal resonances (45,46). Also, in studies across subjects or tracking a single subject over time, differing volume compositions could obfuscate any metabolic changes. In that case, careful positioning of the volume to match compositions should be done using a tool such as ImScribe (https://www.med.upenn.edu/CAMIPM/imscribe.html) that coregisters a high resolution anatomic image to a previously-acquired template and returns the location and orientation of the matching spectroscopy volume.

In addition, while the low bandwidth refocusing pulses deliberately exploit the CSDA of the water and lipid signals, undesired CSDA is also elicited over the excited spectral range of interest. For NAD^+^, the CSDA between the undetected H5 proton at 8.2 ppm and its coupled partners, the H4 and H6 protons, leads to anomalous J-modulation of those peaks which could affect quantitation. For the protocols described here, the minimum TE is much less than 1/J (19); therefore, the effect on quantitation should be mild and could be taken into account using prior-knowledge based fitting. Nevertheless, this technique of low bandwidth localization is best suited when the spectral range of interest is small compared to its spectral separation from nuisance signals.

## CONCLUSION

This study describes ^1^H spectroscopic techniques specifically targeting low concentration metabolites with downfield resonances using large volume acquisitions. Utilizing novel acquisition and processing methods, we have achieved improved spectral quality at 7T in brain and calf muscle as well as unmatched spectral quality in brain at 3T, detecting whole brain NAD^+^ and tryptophan in under 5 minutes.

## Data Availability

All data produced in the present study are available upon reasonable request to the authors.

---

**Supporting Information Figure S1:**
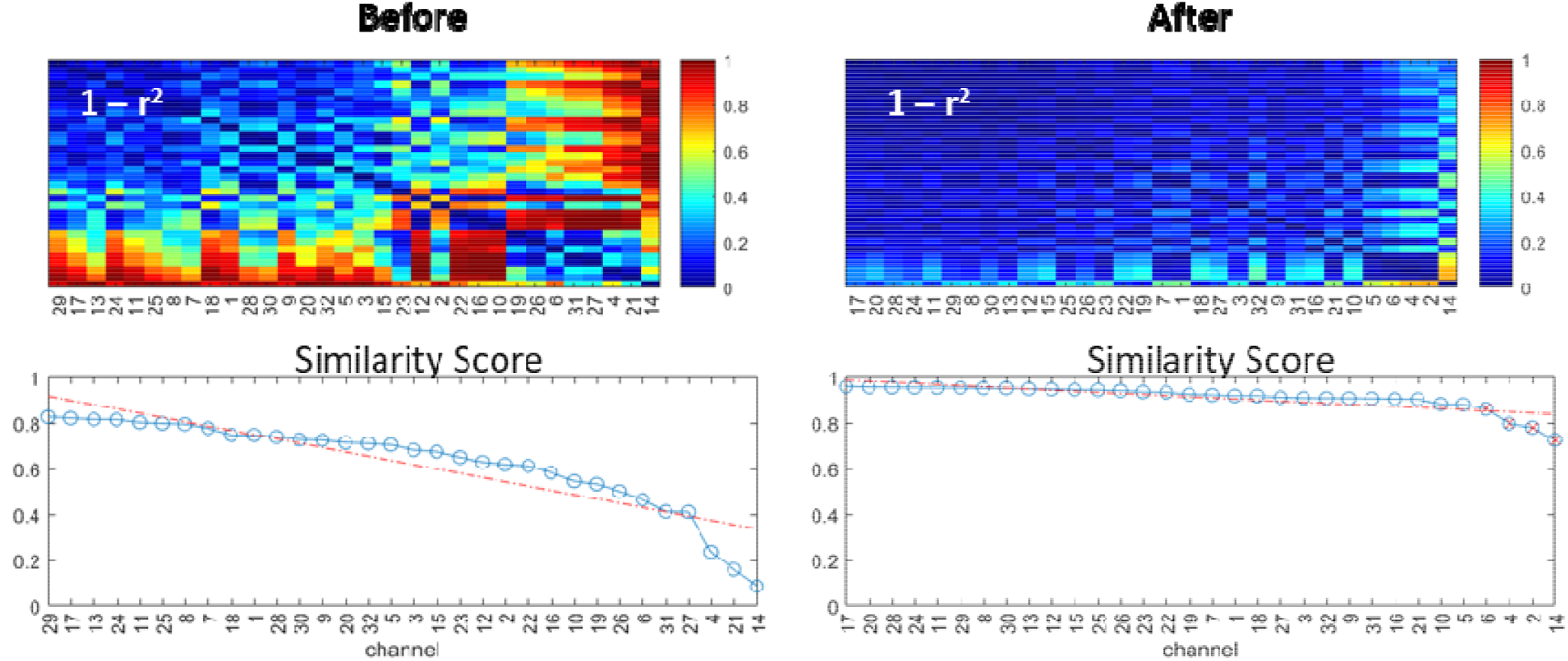
Between-channel correlation matrices (top row) and ranked channel-wise similarity score plots (bottom row), computed before (left) and after (right) channel-wise frequency alignment of a water reference scan. The lowest ranked channels (red x), whose similarity scores fall below the dashed red line, are pruned from the coil combination step.

**Supporting Information Figure S2:**
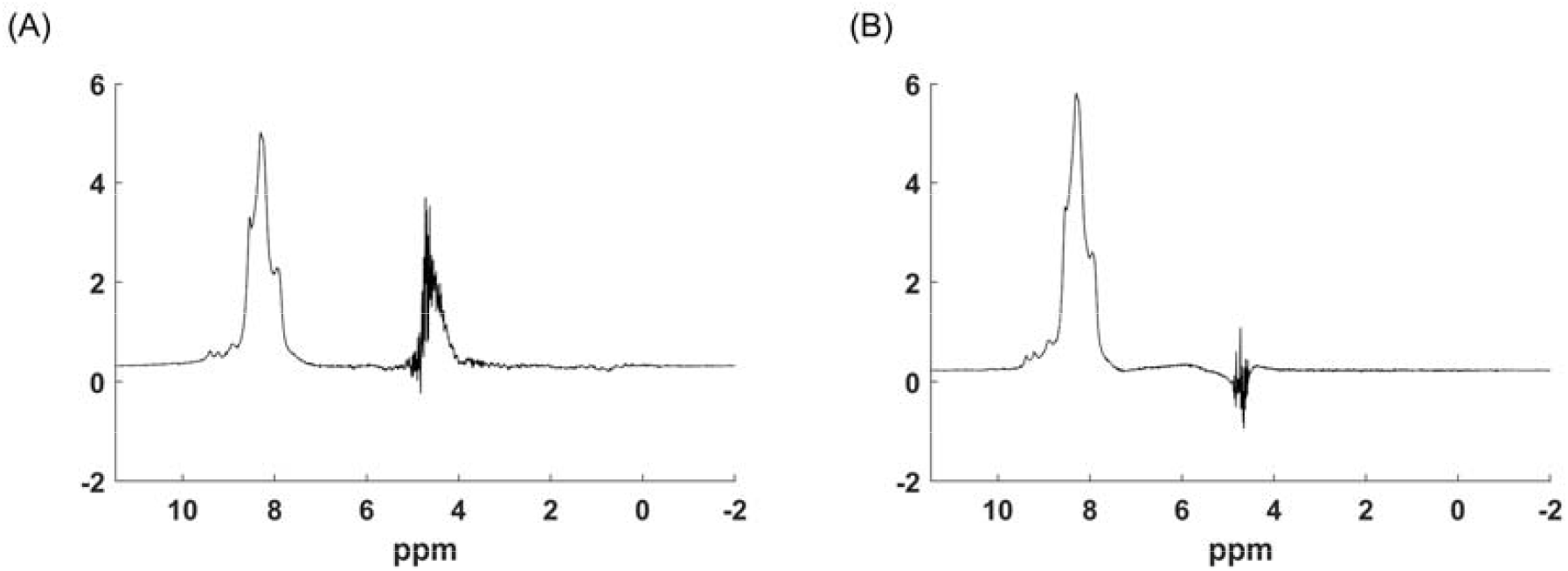
Comparison of spectra acquired in brain from the same slice in the same session with only the direction of the CSDA reversed. The water volume is shifted (A) inferiorly or (B) superiorly. Because of the low B1+ of the 1 Tx/64 Rx head coil at the base of the brain, low water signal is observed even with an inferior CSDA, even though the differences in water signal at each location are vast.

**Supporting Information Figure S3:**
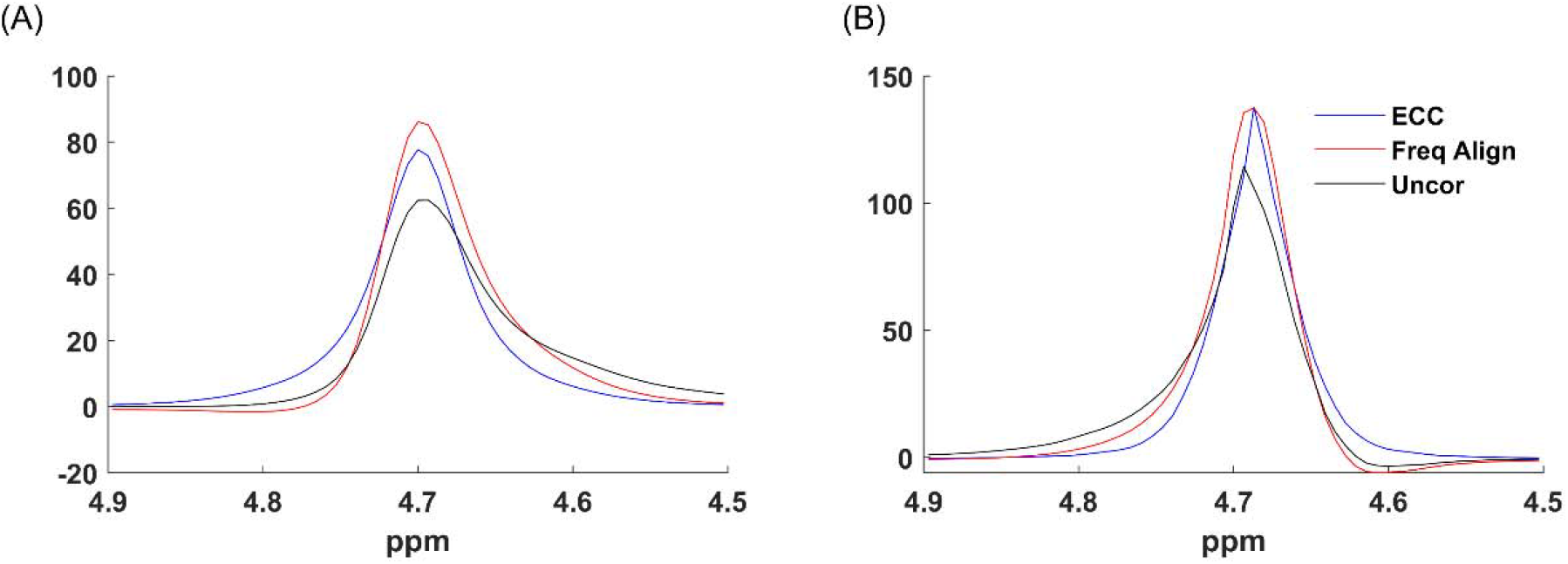
Channel-wise corrections of a water reference scan. Uncorrected (black), eddy current corrected (blue), and frequency aligned and pruned (red) for a dataset showing the relative improvement of the proposed method compared to eddy current correction (A) as well as a dataset showing little difference between the two (B). Both corrections show improved peak shape and linewidth compared to the uncorrected signal. All peaks in each subfigure are normalized to have the same area.

